# Could the new COVID-19 mutant strain undermine vaccination efforts? A mathematical modelling approach for estimating the spread of the UK mutant strain using Ontario, Canada, as a case study

**DOI:** 10.1101/2021.02.02.21251039

**Authors:** Matthew Betti, Nicola Luigi Bragazzi, Jane Marie Heffernan, Jude Kong, Angie Raad

## Abstract

**Background:** Infections represent highly dynamic processes, characterized by evolutionary changes and events that involve both the pathogen and the host. Among infectious agents, viruses, such as the “Severe Acute Respiratory Syndrome-related Coronavirus type 2” (SARS-CoV-2), the infectious agent responsible for the currently ongoing “Coronavirus disease 2019” (COVID-2019) pandemic, have a particularly high mutation rate. Taking into account the mutational landscape of an infectious agent, it is important to shed light on its evolution capability over time. As new, more infectious strains of COVID-19 emerge around the world, it is imperative to estimate when these new strains may overtake the wild-type strain in different populations. Therefore, we developed a general-purpose framework to estimate the time at which a mutant variant is able to takeover a wild-type strain during an emerging infectious diseases outbreak. In this study, we used COVID-19 as a case-study, but the model is adaptable to any emerging pathogens.

**Methods and findings:** We devise a two-strain mathematical framework, to model a wild- and a mutant-type viral population and fit cumulative case data to parameterize the model, using Ontario as a case study. We found that, in the context of under-reporting and the current case levels, a variant strain is unlikely to dominate until March/April 2021. Current non-pharmaceutical interventions in Ontario need to be kept in place longer even with vaccination in order to prevent another outbreak. The spread of a variant strain in Ontario will mostly likely be observed by a widened peak of the daily reported cases. If vaccine efficacy is maintained across strains, then it is still possible to have an immune population by end of 2021.

**Conclusions:** Our findings have important practical implications in terms of public health as policy-and decision-makers are equipped with a mathematical tool that can enable the estimation of the take-over of a mutant strain of an emerging infectious disease.

## 1 Introduction

Infections represent highly dynamic processes, characterized by evolutionary changes and events that involve both the pathogen and the host [1] and can be understood at two levels, namely the intra- and the inter-host levels [2]. Among infectious agents, viruses have a particularly high mutation rate, which is even more relevant in terms of public health control and management considering their short generation times and relatively large population sizes [1]. It is of paramount importance to take into account the mutational landscape of an infectious agent, to shed light on its evolution capability over time, to be able to capture events leading to a rapid and effective adaptation to the host environment, impacting its fitness and transmissibility to new hosts [3].

Pathogen evolution and recombination can result in escaping the host immune system, causing drug failure and leading to the insurgence of anti-microbial drug resistance. Further, it can compromise the effectiveness of existing vaccines making infection prevention and control more challenging [3].

The “Severe Acute Respiratory Syndrome-related Coronavirus type 2” (SARS-CoV-2) is the infectious agent responsible for the currently ongoing “Coronavirus disease 2019” (COVID-2019) pandemic. COVID-19 is a generally mild but sometimes severe and even life-threatening communicable disease [4]. This novel, emerging coronavirus exhibits a constantly and dynamically evolving mutational landscape, with a relatively abundant genetic diversity [5] and a high evolution capability over time [6]

As new, more infectious strains of COVID-19 emerge around the world [7], it is imperative to estimate when these new strains may overtake the wild-type strain in different populations. Therefore, we developed a general-purpose framework for estimating takeover of mutant strains of emerging infectious diseases. In the this study, we used COVID-19 as a case-study, but the model is also adapted to any emerging pathogen.

## 2 Methods

We extend the model in [8]. We maintain a wild-type population and fit cumulative case data to parameterize the wild-type model. We simplify the model to take into account total cases *C*_*I*_, known cases *C*_*K*_, mild active cases *I*_*m*_ and active severe cases *I*_*s*_. The wild-type model equations first presented in [8] is modelled by the below equations:

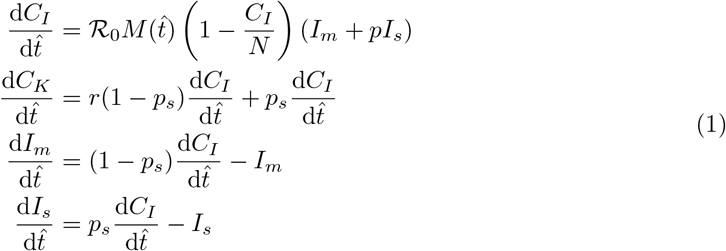

where 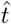 is time in the units of infectious lifetime, ℛ_0_ is the basic reproduction number, *M* (*t*) is a mitigation function that describes non-pharmaceutical interventions, *N* is the population of the region, *p* is the relative infectiousness of severe cases to mild cases, *r* is the average reporting rate of mild cases, and *p*_*s*_ is the probability that a case is severe.

The model comes with a set of assumptions that are discussed in [8], highlighted below:

1. Reporting is relatively consistent.
2. The total population in a region is constant.
3. All severe cases are reported.

We note that second assumption implies that the model is for short-term projection and third assumption implies that severe cases will always require medical intervention and is thus always reported. We also point out that first assumption and by virtue of the model itself, this model assumes that an outbreak is mainly being driven by community transmission.

Using a least square method, we fit *C*_*K*_ to reported cumulative case data in the province of Ontario and get estimates for total cases and active mild/severe cases.

With base parameters we can then extend the model to account for a more infectious variant. We still fit to the same known infections, *C*_*K*_, but we require the wild-type given by *I*_*m*_ and *I*_*s*_ as well as a mutant strain *Ĩ* _*m*_ and *Ĩ* _*s*_. The full model with both strains is then given by

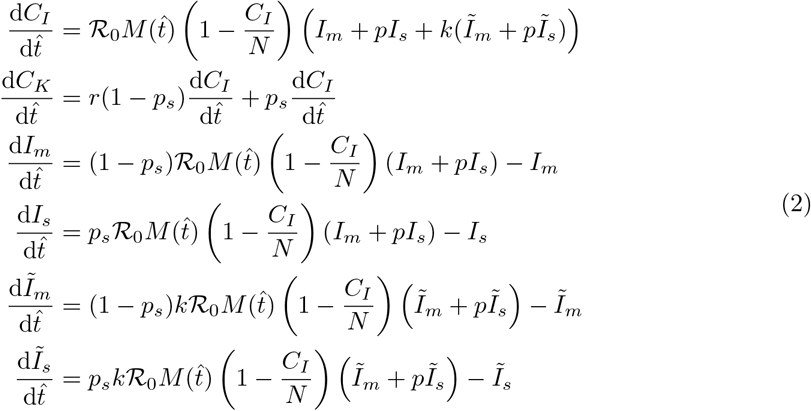

## 3 Results

### Scenario-based extrapolation from first known cases of UK-variant in Ontario

#### No vaccination, no relaxation

In Figure 1, we extrapolate fits from December 16 2020 to January 11 2021 to account for the presence of the COVID-19 UK variant in Ontario where two confirmed cases were reported on December 26, 2020. Using the fitted average reporting rate, we assume that there are roughly 60 active cases of the variant in the province at the time and that the variant is 1.7 times as infectious as the wild-type strain REFXXX. We see that if introduced around Christmas time, and non-pharmaceutical interventions are continued to tighten, the mutant strain cannot saturate until late 2021 and will not become the dominant strain until April 2021, as seen in Figure 2. This scenario acts as a control as it does not account for vaccination, or relaxation of NPIs.

**Figure 1:**
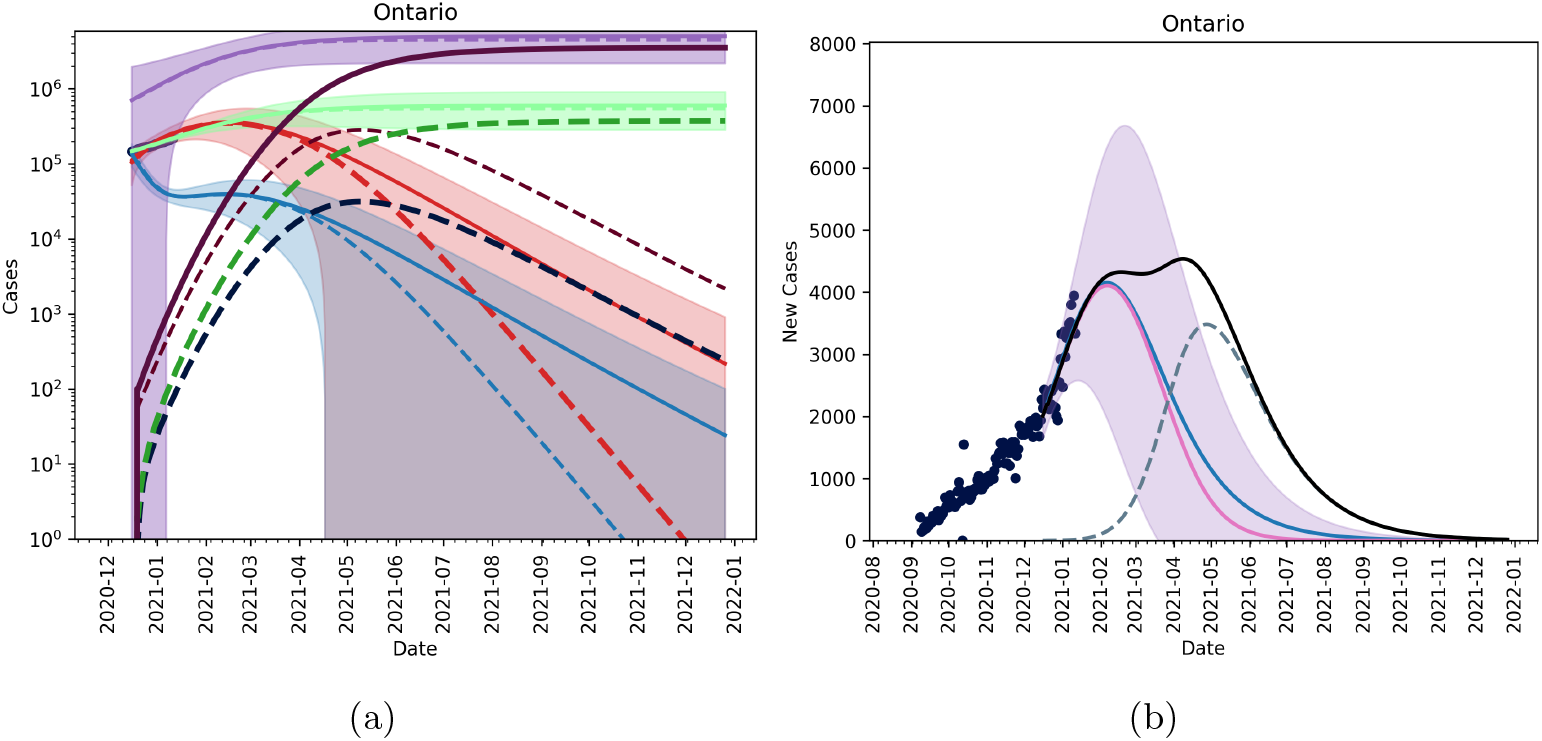
Figures showing the fitted trajectory against the introduction of a mutant strain. Subfigure (a) shows active and cumulative cases for model without a mutant (solid lines) and model with mutant strain (dashed lines). Red is the active mild cases of the wild type, blue is the active severe cases of the wild type, purple is the cumulative wild type cases and light green is the cumulative reported cases of the wild time. Dark red, dark blue, dark purple and dark green are active mild, active severe, cumulative and reported cumulative cases for the mutant strain. Subfigure (b) shows new reported cases per day with blue being the fitted data with no mutant introduction, pink is the wild-type, and dashed blue is the mutant strain. The black line shows total new cases per day. We assumed 60 active cases on December 26, 2020.

**Figure 2:**
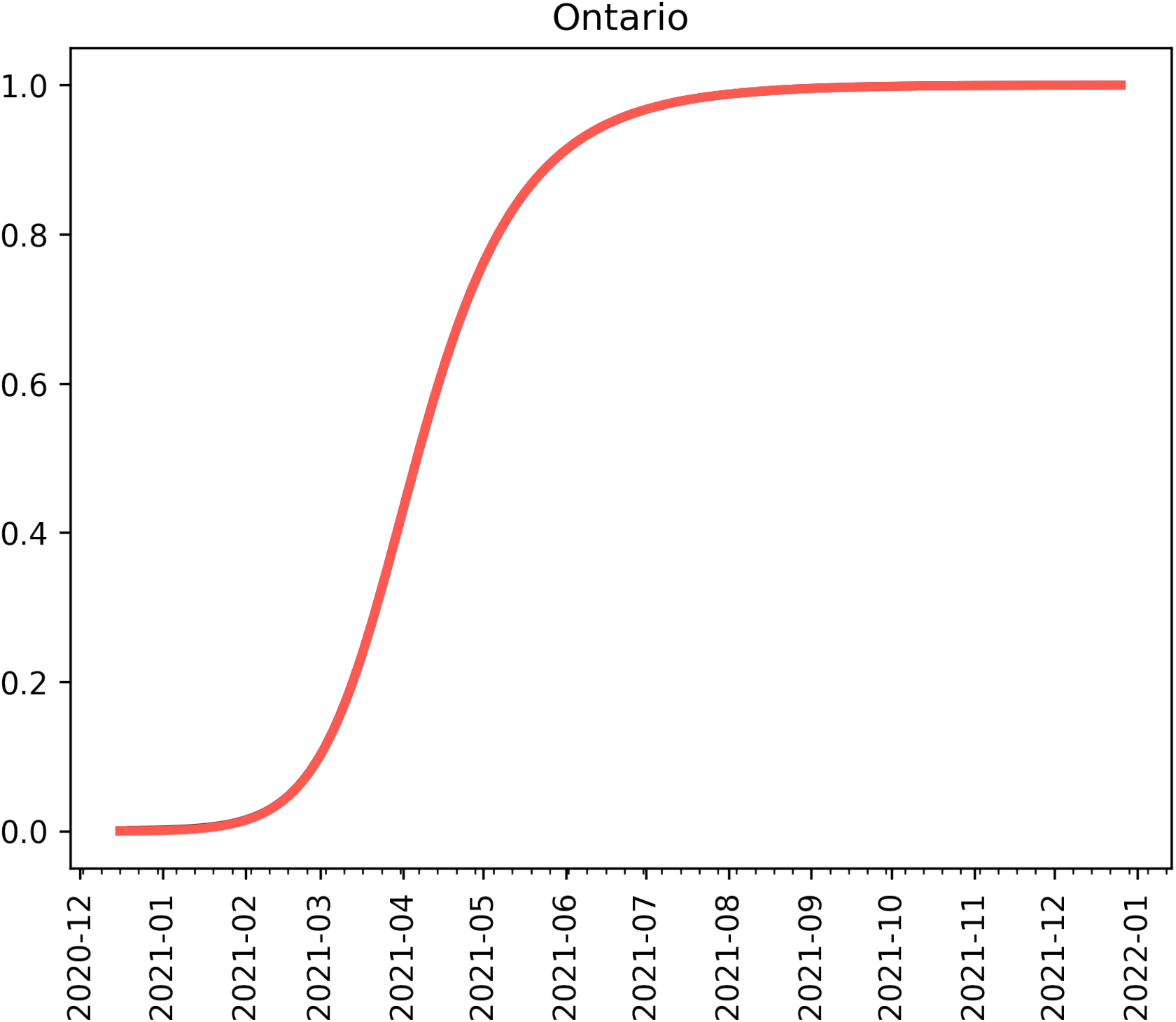
Proportion of active cases which are the mutant assuming 60 active cases on December 26, 2020.

In Figure 3 we see that if we assume that there are many cases of the UK-strain in Ontario (1000 cumulative cases as of December 26, 2020) by the time we are able to detect 2 cases, then it is possible for the mutant strain of the SARS-CoV-2 virus to become the dominant strain by March/April 2021 under the current NPIs measure in place. Figure 4 shows that by early March the new mutant will account for the majority of active cases in the province of Ontario.

**Figure 3:**
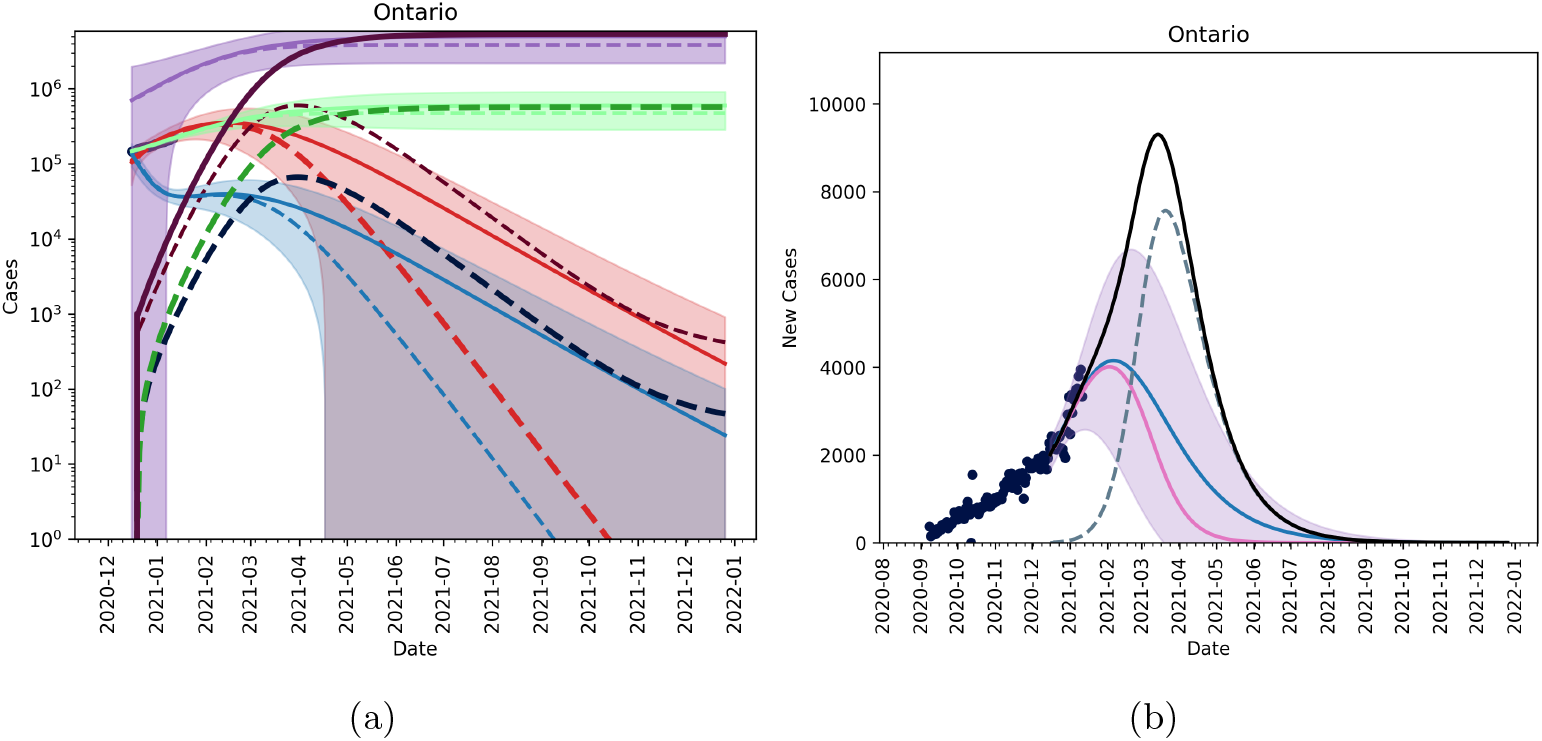
Figures showing the fitted trajectory against the introduction of a mutant strain. Subfigure (a) shows active and cumulative cases for model without a mutant (solid lines) and model with mutant strain (dashed lines). Red is the active mild cases of the wild type, blue is the active severe cases of the wild type, purple is the cumulative wild-type cases and light green is the cumulative reported cases of the wild time. Dark red, dark blue, dark purple and dark green are active mild, active severe, cumulative and reported cumulative cases for the mutant strain. Subfigure (b) shows new reported cases per day with blue being the fitted data with no mutant introduction, pink is the wild-type, and dashed blue is the mutant strain. The black line shows total new cases per day. We assumed 600 active cases on December 26, 2020.

**Figure 4:**
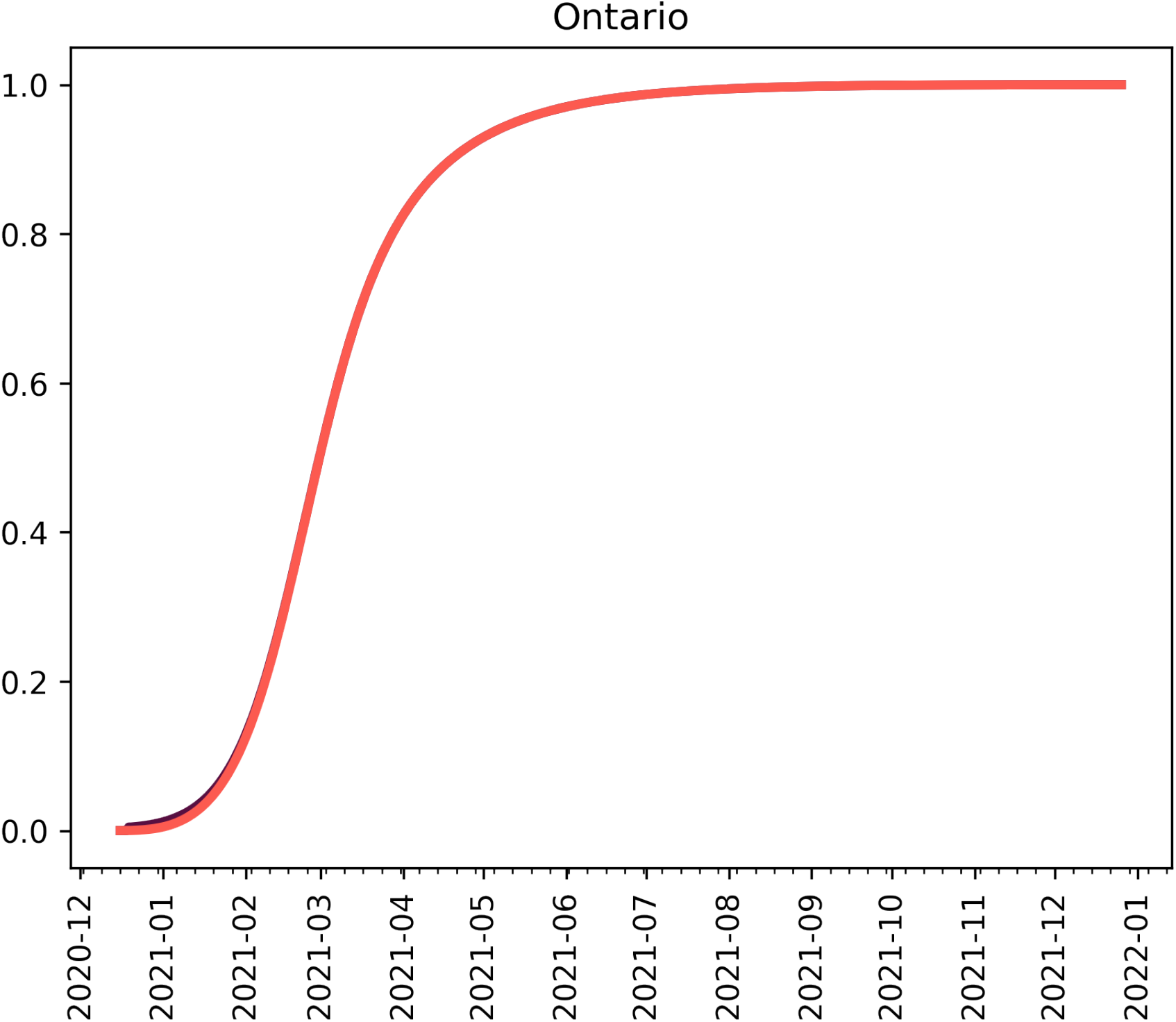
Proportion of active cases which are the mutant assuming 600 active cases on December 26, 2020.

**Figure 5:**
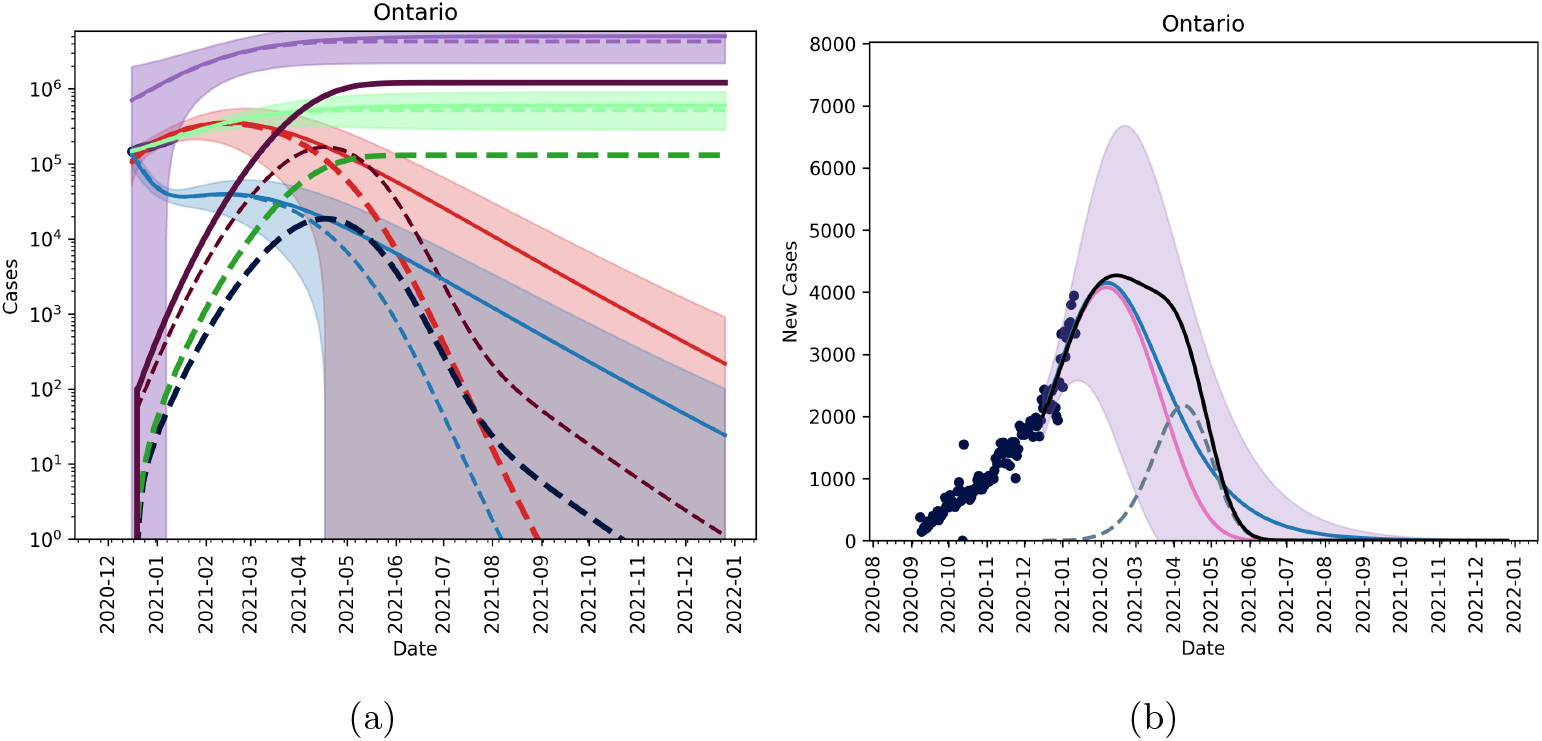
Figures showing the fitted trajectory against the introduction of a mutant strain. Subfigue (a) shows active and cumulative cases for model without a mutant (solid lines) and model with mutant strain (dashed lines). Red is the active mild cases of the wild type, blue is the active severe cases of the wild type, purple is the cumulative wild type cases and light green is the cumulative reported cases of the wild time. Dark red, dark blue, dark purple and dark green are active mild, active severe, cumulative and reported cumulative cases for the mutant strain. Subfigure (b) shows new reported cases per day with blue being the fitted data with no mutant introduction, pink is the wild-type, and dashed blue is the mutant strain. The black line shows total new cases per day. We assume that 10% of the population is vaccinated by March 31, 2021 and that 75% of the population inoculated by the end of 2021.

**Figure 6:**
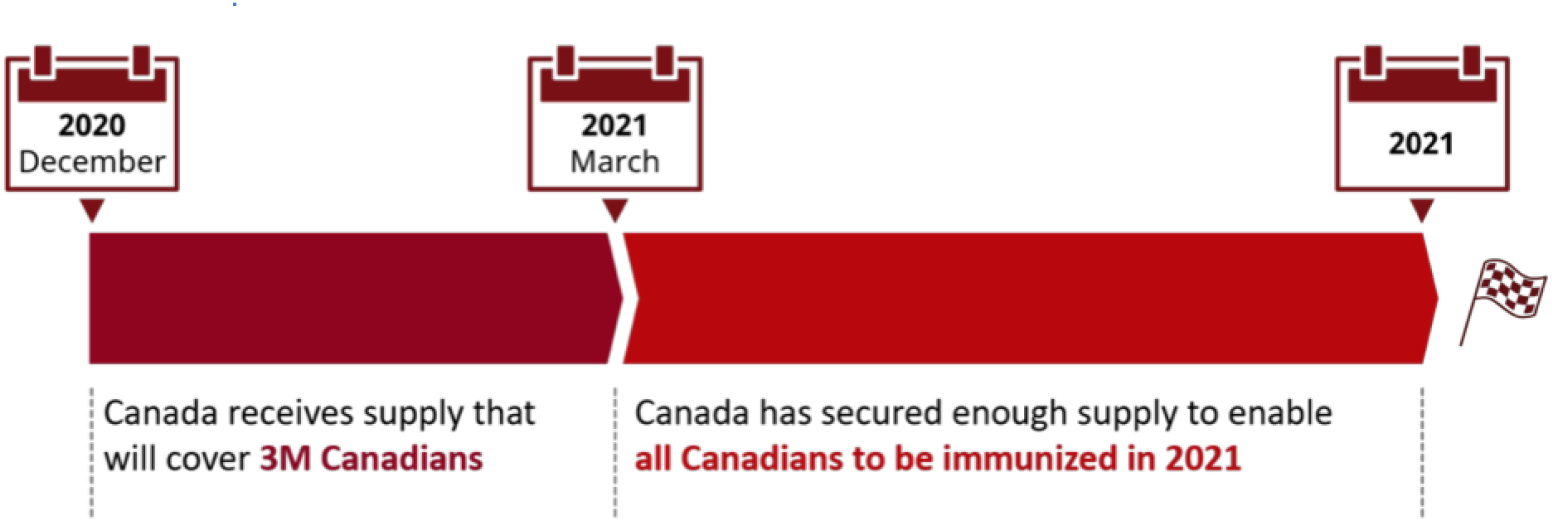
The Federal Government vaccine roll-out plan, adapted from [10].

#### Vaccination without relaxation

Figure (5) shows that a fairly aggressive vaccination plan, namely the vaccination plan outlined in Figure (6) where 10% of the population is vaccinated by second quarter of 2021 and 75% of the population is vaccinated by the end of 2021. We observe that compared to the results in [9], the new, more infectious strain will require NPIs to be in place much longer. If we maintain and escalate NPIs such that we can reduce transmission by 60% we can still lower cases to near zero by July/August 2021. We can compare this to the results in [9] where as we only required a 20% reduction in transmission through NPIs, plus vaccination to see a similar effect.

#### Vaccination and relaxation

Figure 7 shows that slow, controlled relaxation starting May 2021 will result in a small outbreak of the UK variant. In Figure 8 (a) and (b) we see that relaxing immediately on May 1 (i.e. the basic reproduction number of the wild type goes back to *R*_0_ *≈* 2.5), will result in the emergence of a new outbreak, mainly dominated by the new strain.

**Figure 7:**
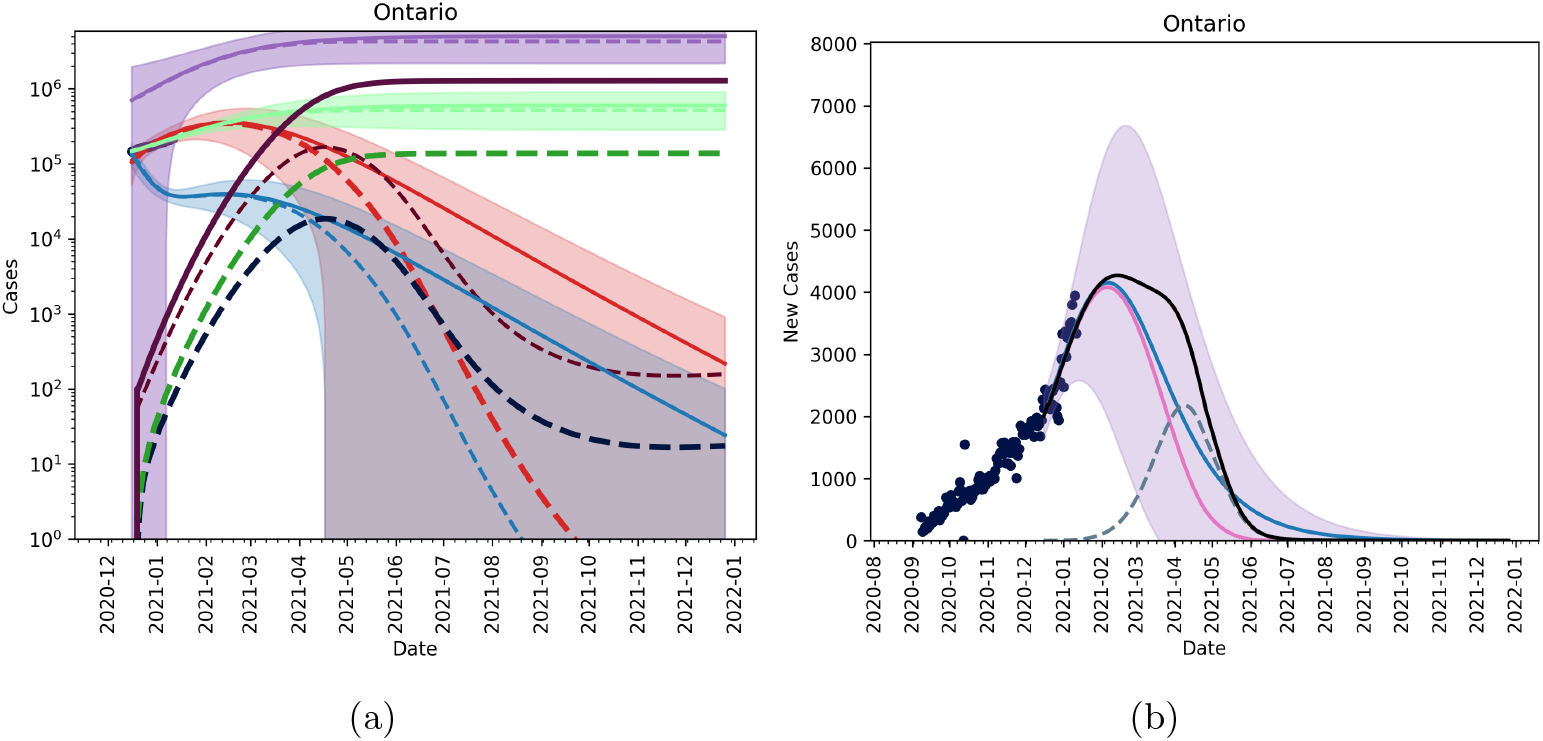
Figures showing the fitted trajectory against the introduction of a mutant strain. Subfigure (a) shows active and cumulative cases for model without a mutant (solid lines) and model with mutant strain (dashed lines). Red is the active mild cases of the wild type, blue is the active severe cases of the wild type, purple is the cumulative wild type cases and light green is the cumulative reported cases of the wild time. Dark red, dark blue, dark purple and dark green are active mild, active severe, cumulative and reported cumulative cases for the mutant strain. Subfigure (b) shows new reported cases per day with blue being the fitted data with no mutant introduction, pink is the wild-type, and dashed blue is the mutant strain. The black line shows total new cases per day. We assume that 10% of the population is vaccinated by March 31, 2021 and that we approach 75% of the population inoculated by the end of 2021 and that NPIs are slowly relaxed starting on May 1, 2021 through the summer.

**Figure 8:**
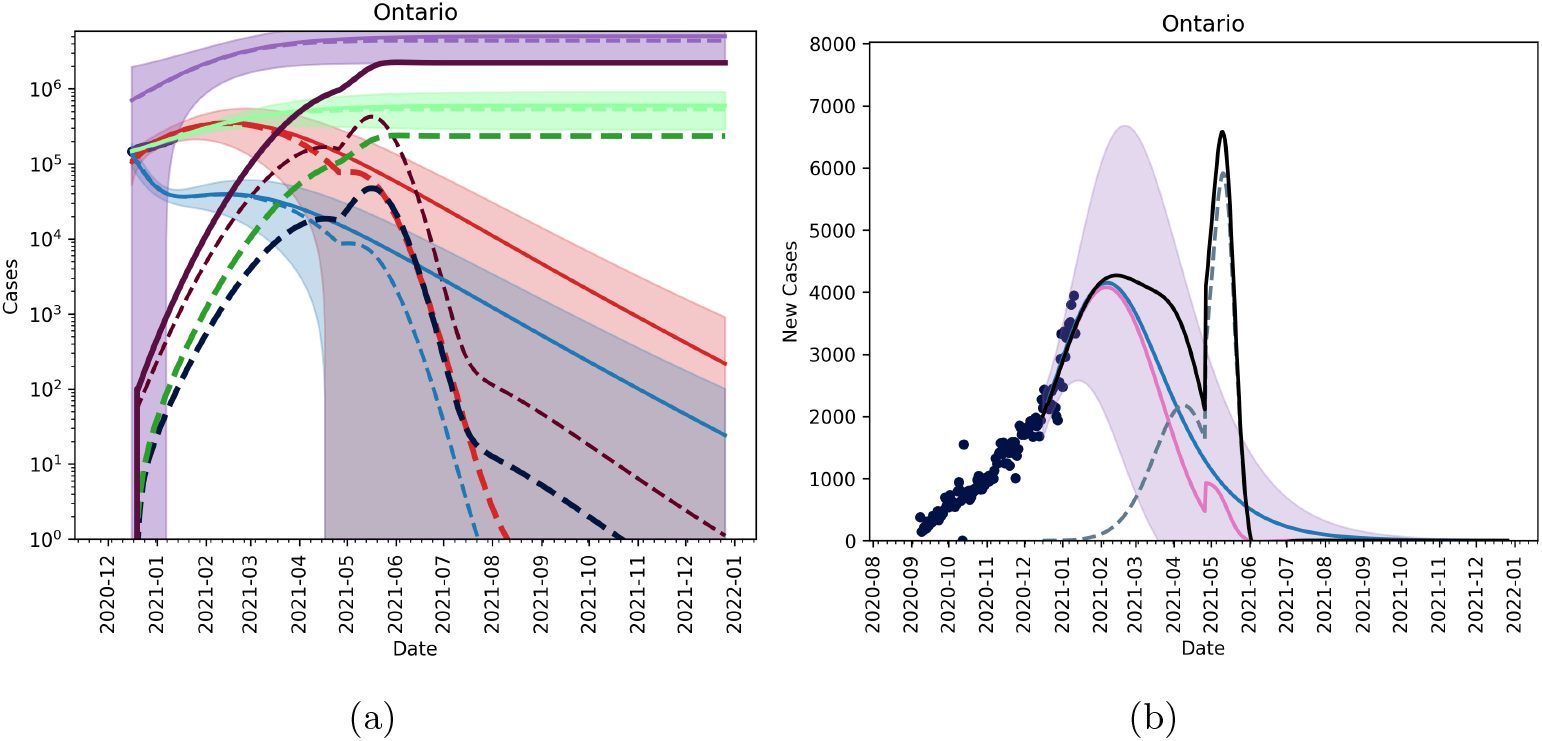
Figures showing the fitted trajectory against the introduction of a mutant strain. Subfigure (a) shows active and cumulative cases for model without a mutant (solid lines) and model with mutant strain (dashed lines). Red is the active mild cases of the wild type, blue is the active severe cases of the wild type, purple is the cumulative wild type cases and light green is the cumulative reported cases of the wild time. Dark red, dark blue, dark purple and dark green are active mild, active severe, cumulative and reported cumulative cases for the mutant strain. Subfigure (b) shows new reported cases per day with blue being the fitted data with no mutant introduction, pink is the wild-type, and dashed blue is the mutant strain. The black line shows total new cases per day. We assume that 10% of the population is vaccinated by March 31, 2021 and that we approach 75% of the population inoculated by the end of 2021 and that NPIs are slowly relaxed immediately on May 1, 2021.

#### Christmas as an anomaly

We can use fits from September 9, 2020 up to December 9, 2020 instead of our more recent fits. This approach will treat the weeks around Christmas as an anomaly. A posteriori, we can see that this indeed likely as caseloads have fallen again in January. Using these fits, along with our candidate vaccination strategy and May 1 relaxation we see that the proliferation of a new, more infectious strain is likely to create a prolonged ‘peak’ new daily infectious. However, if the vaccine is effective against the new strain, the time to herd immunity remains largely unchanged. This is shown in Figure 9.

**Figure 9:**
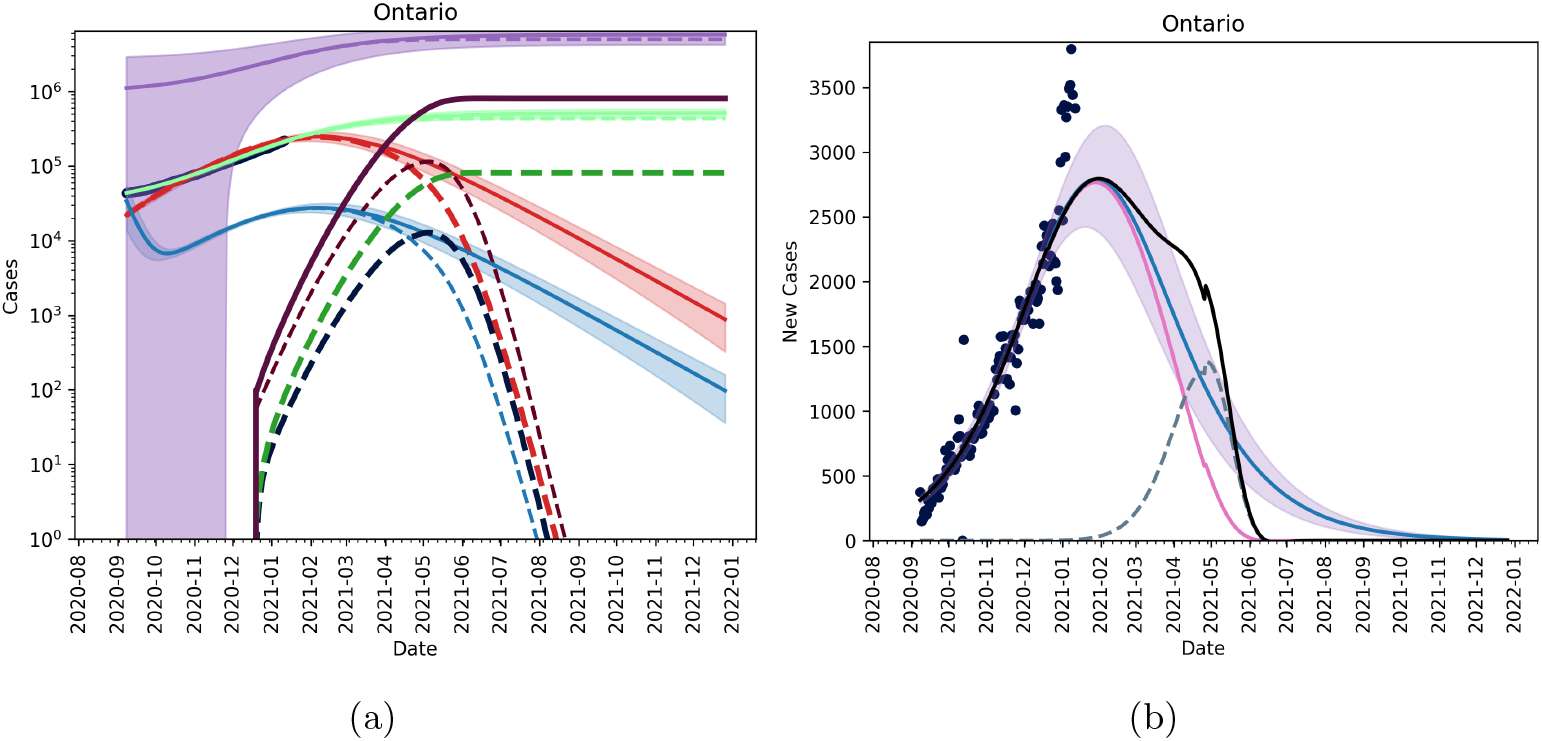
Figures showing the fitted trajectory against the introduction of a mutant strain treating the weeks around Christmas as an anomaly. Subfigure (a) shows active and cumulative cases for model without a mutant (solid lines) and model with mutant strain (dashed lines). Red is the active mild cases of the wild type, blue is the active severe cases of the wild type, purple is the cumulative wild type cases and light green is the cumulative reported cases of the wild time. Dark red, dark blue, dark purple and dark green are active mild, active severe, cumulative and reported cumulative cases for the mutant strain. Subfigure (b) shows new reported cases per day with blue being the fitted data with no mutant introduction, pink is the wild type, and dashed blue is the mutant strain. The black line shows total new cases per day. We assume that 10% of the population is vaccinated by March 31, 2021 and that we approach 75% of the population inoculated by the end of 2021 and that NPIs are slowly relaxed starting on May 1, 2021 through the summer.

## 4 Discussion

SARS-CoV-2 is an emerging coronavirus responsible for the still ongoing COVID-19 pandemic. Ontario, Canada, as well as other territories and countries worldwide have experienced multiple waves, and have been struggling to find a difficult compromise between, on the one hand, ensuring and guaranteeing safety and, on the other hand, preserving at least essential businesses by modulating the stringency of public health interventions, strengthening/relaxing them based on up-to-date epidemiological data. One year after the initial outbreak that emerged in December 2019, a number of vaccines have been approved, even though new variants characterized by higher transmissiblity have been detected.

Like other viruses causing widespread transmission in the population, SARS-CoV-2 has mutated many times since its initial insurgence (an outbreak of pneumonia of unknown etiology occurred in Wuhan, province of Hubei, mainland China). Based on its genomic profile, SARS-CoV-2 can be subdivided into various genetic groups, known as clades. A set of specific mutations would enable researchers to distinguish between viral groups currently dominating and circulating worldwide. These groups are generally called lineages, even though the precise nomenclature and the taxonomic hierarchy of SARS-CoV-2 are still under debate and, generally speaking, classifying viral variety and diversity is a rather challenging task [11, 12].

Mutations arise spontaneously as a consequence of a complex, multi-factorial series of macro- and micro-evolution processes as well as the result of selection pressures [7]. However, some of these mutations (termed as “variants of concern”, VOCs) may be particularly clinically meaningful, especially from the public health perspective, being associated with higher force of infection, transmissibility as well as mortality [7]. In particular, since December 2020, some VOCs have been reported by national public health authorities to the World Health Organization (WHO), including VOC-202012/01 (also known as lineage B.1.1.7, commonly referred to as the “UK variant” or the “British variant”), 20I/501Y.V2 (known as lineage B.1.351, commonly termed as the “South African variant”) and lineage B.1.1.28 (known as the “Brazilian variant”). Other variants are under investigation and strict follow-up from international public health bodies, including the “Japanese variant” (variant P.1, lineage B.1.1.28) and the “USA variant” (L452R). This topic is constantly under flux as identifying the impact of a variant is of paramount importance. Once introduced in the population, a highly transmissible variant could become more and more prevalent, leading to the replacement of the original wild strain and making infection control and management particularly difficult.

Our findings have important practical implications in terms of public health as policy- and decision-makers are equipped with a mathematical tool enabling the estimation of the take-over of a mutation strain of an emerging infectious disease, such as the previously mentioned VOCs. Moreover, in this paper, we identified that, in the context of under-reporting and the current case levels, a variant strain is unlikely to dominate until March/April 2021. Current NPIs in Ontario need to be kept in place for longer even with vaccination in order to prevent another outbreak. The proliferation of a variant strain in Ontario will mostly likely be observed by a widened peak of reported daily cases. If vaccine efficacy is maintained across strains, then it is still possible to have an immune population by end of 2021. A limitation of this model is that it does not account for importation of cases which could prolong outbreaks. With new rules in place by the Government of Canada surrounding international travel, the practical effects of importation are low.

## Data Availability

Data publicly available
https://www.canada.ca/en/public-health/services/diseases/2019-novel-coronavirus-infection/symptoms.html

